# Barriers to early initiation of breastfeeding in healthy neonates in an urban hospital setting

**DOI:** 10.1101/2021.03.19.21253932

**Authors:** Davis Rubagumya, Muzdalfat Abeid, Eric Aghan, Mariam Noorani

## Abstract

**Background:** Breastfeeding is a key intervention to improve global targets on nutrition, health and survival. The World Health Organization (WHO) recommends early initiation of breastfeeding to prevent infections in the newborn and achieve targets of exclusive breastfeeding for the first six months of life. Early initiation of breastfeeding improves neonatal-maternal bonding, reduces jaundice and prevents gastrointestinal and respiratory tract infections. Global prevalence of early initiation of breastfeeding is 45% and 51% for Tanzania. The recommended prevalence is at least 80% by WHO guidelines.

**Objective:** To explore barriers to early initiation of breastfeeding in a hospital setting.

**Methods:** A descriptive exploratory qualitative study with semi-structured individual interviews was employed to explore the barriers to early initiation of breastfeeding in a hospital setting. Three midwives and six mothers were interviewed. The interview topic guide covered experiences and challenges of early initiation of breastfeeding. Data was analyzed using Systematic Text Condensation as described by Malterud.

**Results:** Participants perceived that inadequate breastfeeding information especially on ideal time to start breastfeeding contributed to delayed initiation. The practices and environment post-delivery such as perineal tear repair and dirty labor room prevented women from initiating breastfeeding immediately.

**Conclusions:** The barriers identified were related to gaps in knowledge, immediate postpartum practices and individual perceptions of a non-conducive environment.

## BACKGROUND

Breastfeeding is the biological norm for mammals to provide adequate, pollution free and sustainable nutrition (1). For humans, breastfeeding is important for achieving global targets on nutrition, health, survival, economic growth and environmental sustainability (1). The World Health Organization (WHO) advocates on initiating breastfeeding within the first hour post-delivery and to be practiced exclusively for six months (1). Breastfeeding has benefits for both mothers and newborns. Maternal benefits include uterine contraction hence decreasing postpartum bleeding, decreased risk of depression, increased bonding with the newborn, decreased risks of breast and ovarian cancers, decreased risk of diabetes mellitus type 2 and hypertension (3,4). For newborns, breastfeeding helps in preventing infections especially gastrointestinal and respiratory, offers immune protection hence decreasing the incidences of allergies and celiac disease, enhances neurodevelopment and reduces the risks of obesity (3,4).

Colostrum, which is the first milk produced after delivery has been recommended by WHO as the perfect source of nutrition for newborns (2). Despite having an immature digestive system, colostrum is well tolerated by newborns providing nutrients such as epidermal growth factor that aids in intestinal maturation and repair, help in clearing excess bilirubin, helping in prevention of infections as well as aiding in neurodevelopment (5).

The critical window for initiating lactation is within the first hour after delivery (2). Delayed breastfeeding initiation has been associated with increased risk of cough and difficulty breathing during the first 6 months of life (6). It has been shown that neonatal mortality risk increases linearly with the time of breastfeeding initiation; children who started breastfeeding between 2-23 hours post birth had 33% greater risk of mortality as compared to those whom breastfeeding was initiated within the first hour post-delivery. This risk was 100% greater in infants who were fed after 24 hours post-delivery (7). Early initiation of breastfeeding exposes the newborn to maternal skin flora which help in colonizing the infant’s gut and skin and avoids colonization by harmful bacterial from the care givers and the surrounding (8). Breastfeeding within the first hour also leads to stability of the newborn heart rate and breathing (9).

Globally, 45% of newborns start breastfeeding within the first hour after being born (10). In Tanzania, 51% of newborns start breastfeeding within one hour of birth. This ranges from 26% in Simiyu region to 80% in Tanga region (11). In Dar es Salaam, the percentage of early initiation has been reported to be 51.1% (11). In addition, 14% of ever breastfed infants started with pre-lacteal feeds before initiating breastfeeding despite being contrary to recommendations of exclusive breastfeeding as per WHO guidelines (11).

Several studies have identified factors influencing early initiation of breastfeeding. Some of them are:

### PLACE OF DELIVERY

Institutional deliveries have been shown to facilitate early initiation of breastfeeding. In a cross-sectional survey of random households in rural Tanzania, delivery in a health facility was associated with more women initiating breastfeeding within one hour as compared to those who gave birth at home [53.9% vs 42.3%, OR = 1.75, 95 % CI 1.25, 2.45] (12).

A systematic review has shown that adherence to the Baby-friendly Hospital Initiative (BHFI) increases the rate of early initiation of breastfeeding (14). The BFHI is an initiative by UNICEF and WHO that gives ten steps for successful breastfeeding which describe the role of healthcare workers as well as mothers for preparation and initiation of breastfeeding (15).

In a report from the document ‘CAPTURE THE MOMENT’ by WHO and UNICEF, Rwanda reported an increase of early initiation of breastfeeding from 64% in 2005 to 81% in 2014 because of implementing an intensive campaign advocating breastfeeding practice together with BFHI (13).

### EDUCATION

Mothers’ level of education has been found to impact initiation of breastfeeding. A prospective cross sectional analytical study done by Mahnaz et al showed that women with education below university level were more likely to initiate early breastfeeding (9). However, another study from Nepal which had data from the demographic and health surveys revealed that women who had not attended school delayed breastfeeding initiation as compared to those who had formal education. (16). These studies show conflicting findings about the role of formal education in breastfeeding success.

### MOTHERS AGE

Age of the woman who has given birth is another factor that determines early initiation of breastfeeding, especially for new mothers. In Ethiopia, in a study done by Meseret et al, the odds of early initiation of breast feeding were three times higher for women in the age group of 20-24 as compared to 15-19 (18). There was no further explanation from the study, however teenage pregnancies were associated with poor antenatal attendance as well as lack of support, hence chances of delaying in initiating breastfeeding were increased.

### GESTATIONAL AGE AT BIRTH

#### HISTORY OF BREASTFEEDING

Previous experience of breastfeeding has been associated with improved rates of early initiation. In a study done by Noble et al, women who had breastfed previously had more success in early initiation of breastfeeding (17).

#### MODE OF DELIVERY

Early initiation of breastfeeding should be done irrespective of the mode of delivery. However, studies have shown that vaginal delivery is associated with higher rates of early initiation of breastfeeding as compared to cesarean delivery (9). Some reasons cited for the delay in case of caesarean delivery include: mother’s delay in awakening and regaining of cognitive features after general anesthesia (19) and anticipated effects of anesthetic drugs to newborn via breastmilk. Studies have shown that successful skin to skin contact and early initiation of breastfeeding can be done in the operating theatre after cesarean delivery (21). Adequate support for the mother is of paramount importance, especially in holding a newborn to her chest (19) (22) (20).

#### ROOMING IN

This is one of the key steps in the baby friendly hospital initiative where mother and her baby are kept at close proximity and unnecessary separation is minimized. This has been shown to impact positively on the initiation of breastfeeding within the first hour of life (23).

#### PRE-LACTEAL FEED AVAILABILITY

This involves feeding a newborn with something edible before breastfeeding initiation. This has been shown to result in delayed breastfeeding initiation for newborns (23). In the revised baby friendly hospital initiative document by WHO and UNICEF, emphasis is mentioned on implementing measures to reduce the use of pre-lacteal feeds (1).

#### HOSPITAL POLICIES

Hospital policies and health care personnel provide crucial support for mothers to initiate breastfeeding early (13). In the baby friendly hospital initiative, it has been recommended that hospitals should abide to the WHO code of marketing of breastmilk substitutes and health care providers in the antenatal, delivery, newborn care rooms/wards should be able to explain components of the code (1). It has also been recommended that policies that promote breastfeeding should be implemented and this has been shown to improve early breastfeeding initiation practices (17). In Rwanda, they demonstrated that establishing a cadre of well-trained health professionals to support early initiation is of paramount importance for this practice (13). The World Breastfeeding Trend Initiative Report of 2015 showed that Tanzania scored poorly and this was due to poor implementation of BFHI in the health facilities as well as lack of commitment from the Ministry of Health, Community Development, Gender, Elderly and Children (24)

#### MOTHERS HEALTH POST DELIVERY

For mothers who are HIV positive, guidelines have changed over the years and currently according to Tanzania’s infant feeding guidelines, HIV positive mothers are encouraged to initiate breastfeeding early and continue exclusive breastfeeding while being on antiretroviral treatment (ART) (27).

The mother psychological state post-delivery also contributes to initiation of breastfeeding. A study in Nigeria showed that a good psychological state of the mother was associated with the practice of initiating breastfeeding as recommended (28).

#### SOCIO CULTURAL BELIEFS

Cultural beliefs about colostrum impacts early initiation of breastfeeding. In some cultures, colostrum is considered as medicine whereas other consider it useless milk. A qualitative study done amongst the Sukuma community in Korogocho, Kenya revealed that colostrum was considered to be dirty milk hence has to be expressed and thrown out in the first 2 days. This results in delay in initiation of breastfeeding and in provision of needed nutrition to the newborn baby (25). In Tanzania, a qualitative study on exclusive breastfeeding in Kilimanjaro region revealed that women were hesitant to breastfeed as they feared sagging of their breasts (26).

The report on the situation of infant and young child feeding in Tanzania described several obstacles to proper breastfeeding practices. These include: absence of skilled birth attendant, poor implementation of the international code of marketing of breastmilk substitutes, absence of breastfeeding-related training courses for health professionals and poor implementation of BFHI’s steps (27). Some of the measures that have been recommended to improve breastfeeding practices in Tanzania include: Increase in breastfeeding awareness, men involvement, delivery in presence of skilled personnel and implementation of policies relating to breastmilk substitutes (29–32). However, most of these studies focused on practices of exclusive breastfeeding and there is very little data on early initiation. The present study was done to identify barriers to early initiation of breastfeeding by exploring the experiences of mothers and healthcare providers in a hospital setting.

## METHODS

### STUDY DESIGN

This was a hospital based descriptive exploratory qualitative study that involved individual interview of key informants in exploring the barriers to early initiation of breastfeeding among healthy term neonates in an urban hospital setting. Qualitative methods aid in understanding to a phenomenon under a study in a given setting (33). Individual semi-structured interview was suitable for this particular study because of its ability to aid in exploring study topics in-depth as well as being an attractive approach to a family medicine researcher (34). The consolidated criteria for reporting qualitative research (COREQ) checklist was also employed (35).

### STUDY SETTING

The present study was conducted at The Aga Khan Hospital, Dar es salaam (AKHD), in the obstetrics and gynecology department.

AKHD is a private not-for-profit, Joint Commission International (JCI) accredited, currently a 170-bed, tertiary care hospital with highly specialized staff. It follows the BFHI’s recommended practices including: written breastfeeding policy, encouraging rooming in, skin to skin contact practices and pre delivery breastfeeding education. In addition to that, it does not allow marketing of formula milk and the overall breastfeeding initiation rate while in hospital is close to 100%. It provides comprehensive obstetrics care, including antenatal clinics,24 hours’ emergency obstetric services, delivery services, as well as postnatal care.

In the year 2018, from January to December, there were a total of 1265 deliveries, where by 52.96% were vaginal deliveries, with 23.32% and 23.72% being emergency and elective Caesarean section deliveries.

There are 5 single patient delivery rooms, equipped with delivery bed, baby warmer, and other delivery and resuscitation instruments. Midwives are responsible for conducting deliveries, sometimes with the assistance of Resident doctor or Registered medical officer on duty, and in complex cases, consultants are also available. Every woman in labour has one nurse assigned to attend her. Women who come in labour are assessed by Resident doctor or Registered medical officer, and management plan is communicated to nurses, where by for those whose labour has been confirmed, are taken to the labour room. Moreover, presence of one close relative such as husband, mother or in-laws, is also allowed in the delivery room.

During post-natal care, nurses assess and support breastfeeding and formula milk is only given upon prescription by a pediatrician after a full assessment of breastfeeding. In cases where a new-born weighs 4 kilograms or more, his/her care involves 24 hours observation in the neonatal unit for monitoring of blood sugar levels.

### ETHICS AND CONSENT

The present study was conducted according to universally accepted ethical principles and guidelines for conducting health research in Tanzania. Ethical approval was sought from The Ethics Review Committee of AKU (AKU-ERC). Hospital permission to conduct the study was sought from medical director’s office. Informed written consent was obtained from all participants prior to the interview after provision of needed information and adequate time to participants. Confidentiality was observed in the whole process from data collection to management.

### STUDY PARTICIPANTS

The present study involved midwives and mothers who had delivered healthy term neonates vaginally. Mothers who had postpartum complications were excluded. Purposive sampling was used to recruit participants (36) using a convenience strategy where by information was obtained from participants who met the inclusion criteria and were easily accessible during the study period. The intended sample size was 12 participants since large information power was presumed as the scope of breastfeeding was narrowed to early initiation only in the current study (37). Sample size was determined by the saturation level where by lack of emerging new themes beyond three nurses and six mothers concluded the interviews (33).

### DATA COLLECTION

Semi-structured individual interviews were conducted over a period of four weeks from 2^nd^ November 2019 to 3rd December 2019, where a total of nine participants were involved. All interviews for mothers were done within 48 hours of delivery. To avoid compromising care as well as neonates’ on going breastfeeding, the interviews were conducted through inviting midwives during their off days as well as asking mothers to participate when the neonate was asleep plus presence of a relative who was helping the new mother. The interviews were conducted in Swahili which is the national language of Tanzania. The primary researcher conducted the interviews. An interview guide was used with questions covering specific areas such as knowledge on breastfeeding, ideal time for initiating breastfeeding and experiences concerning breastfeeding practices (appendix 1). The interviews were averaging 30minutes long and audio recorded with permissions from participants. The audio files were transcribed and then translated to English by the researcher. The English versions were reviewed by International English Language Testing System’s certified Swahili speaking person for grammatical corrections followed with translation to Swahili for comparisons with original Swahili transcribed version. No significant mismatches were encountered.

### DATA ANALYSIS

The study process was as shown in the diagram above. No software was employed during the analysis process as data were handled manually. As described by Malterud, Systematic Text Condensation was used on the collected data for analysis (38). This method was considered to be sufficient to analyze the manifest of the data’s contents revealing the challenges to initiation of breastfeeding based on experience from mothers and midwives. The steps employed here involved multiple reading and rereading of the English transcripts to uncover the emerging themes and meaning units hence getting the total impression of the collected materials. The sorted meaning units were coded and grouped into categories and subcategories which were stamped at the manifest level and ratified against the original transcripts as shown in table below:

**Table 1.** Analysis.

| Meaning unit | Category | Subcategory |
| --- | --- | --- |
| 'I had a tear that needed to be repaired' | Post-delivery events hinder breastfeeding | Genital tears repair |
| '.. even in delivery room its possible despite some ladies refusing because of the surrounding being shabby sometimes., imagine breastfeeding in place full of blood!!! | Non-conducive breastfeeding environment | Dirty surroundings |

## RESULTS

Nine participants were interviewed individually where by six were mothers who have delivered within 48 hours and three were nurses who work in the labor room at Aga Khan Hospital Dar es Salaam. The characteristics of participants were as shown in table below.

**Table 2:** Baseline characteristics.

| VARIABLE |  | NURSES | MOTHERS |
| --- | --- | --- | --- |
| AGE (years) | 18 – 30 |  | 3 |
|  | 31-40 | 1 | 3 |
|  | 40< | 2 |  |
| PARITY | 1 | 1 | 2 |
|  | 2 | 1 | 4 |
|  | 5 | 1 |  |
| MARITAL STATUS | Married | 3 | 6 |
| OCCUPATION | Employed worker | 3 | 2 |
|  | Trader |  | 4 |
| EDUCATION | Diploma | 2 | 2 |
|  | Degree | 1 | 4 |

The atmosphere of the arena for interview had features of freedom of expression that was at times spiced by smiles and laughter. The highlights of barriers according to experiences were encountered as the talk ensued. Three main categories emerged as far as barriers to early initiation of breastfeeding is concerned.

**Table 3:** Categories and subcategories.

| **CATEGORIES** | **SUBCATEGORIES** |
| --- | --- |
| **Inadequate information on breastfeeding** | Conflicting information from different sources |
|  | Information on breastmilk substitutes |
|  | Inadequate antenatal education |
|  | Unawareness on time for initiation |
| **Immediate post-delivery events hinder breastfeeding** | Genital tears repair |
|  | Routine post-delivery practices |
| **Non conducive breastfeeding environment** | Lack of care provider support |
|  | Mothers readiness to breastfeed |
|  | Male presence |
|  | Dirty surroundings |

## INADEQUATE INFORMATION ON BREASTFEEDING

All participants perceived lack of proper information as a hindrance to early initiation of breastfeeding. They felt that different sources have conflicting information and without guidance from health personnel, initiation of breastfeeding is hampered. Moreover, they felt that inadequate information during their antenatal clinic visits from both nurses and doctors contributed to delay in initiation of breastfeeding after delivery.

### Conflicting information from different sources

participants felt that information received on breastfeeding from various sources was confusing with some advocating the use of breastmilk while others were for formula milk for newborn babies. In addition, information about body changes after breastfeeding were shared within communities and also negatively impacted breastfeeding.

> *‘…In street, there is a notion that when you breastfeed, your breasts lose shape, they become pendulous like pawpaw (laughing), so for {‘sister du’} fashion girl it becomes tough to think about breastfeeding rather formula milk roams in head. (smiling)’ (Mother 2)*
>
> *‘…I feel it’s good to be informed because sometimes you might come with street information and your mind is just thinking about formula’ (Mother 2)*

### Information on breastmilk substitutes

participants spoke vividly on formula milk. They mentioned that not only availability but also abundant information on social media about it as compared to breastmilk does hamper the ability of women to start breastfeeding after giving birth. This resulted in women expecting to receive formula milk for their newborns.

> *‘*.. *I can tell you; a mother will come knowing there is alternative to breastmilk hence won’t bother with breastfeeding. (smiling)’ (Mother 4)*
>
> *‘I hear about formula milk more than breastmilk’ (Mother 6)*

### Inadequate antenatal education

this was reported by participants as one of the major barriers to early initiation of breastfeeding after delivery. They felt that information from the clinic should include detailed breastfeeding information apart from assessing the wellness of the unborn baby. Moreover, it was mentioned that with personal differences, information on breastfeeding measures should be individualized since different people have different body morphology, for instance having flat nipples where some measures have to be taken so as to be able to breastfeed a newborn. There was frustration expressed by midwives whenever a mother is found to have no breastfeeding knowledge after giving birth despite attending clinic on regular basis as recorded on her antenatal card.

> *‘…. hmm! The doctor mostly would enquire about the age of pregnancy, laboratory results, listening to the baby’s heartbeats, but concerning breastfeeding, no information about that’ (Mother 2)*
>
> *‘…You might find a woman has seven visits and has flat nipple, meaning no one informed her even about preparing for breastfeeding despite such a condition’ (Nurse 2)*
>
> *‘…Sometimes it’s frustrating, you find a lady having (‘sifuri’) zero knowledge on breastfeeding and has a documentation of six clinic visits’ (Nurse 3)*

### Unawareness on time for initiation

this was reported differently by participants hence indicating lack of knowledge on the ideal time for breastfeeding among mothers and care givers. One went on to mention that knowing the exact time within which breastfeeding should start would have prompted her to remind care givers incase time is elapsing without helping her to initiate breastfeeding.

> *‘…I believe, immediately after being born’. ‘Means time (for initiation of breastfeeding) should be within three hours, and the mother has to be back in the ward and comfortable. (laughing)’ (Mother 2)*
>
> *‘…I need to know the time, I never knew that, khaa!!’. ‘if you know that immediately means within one hour, reminding a nurse after delivery becomes easy’ (Mother 6)*
>
> *…hmm! I have never heard of such a time. (with confidence), however it is supposed to be immediately, but about time, even in books, that is not there. (Nurse 2)*

## IMMEDIATE POSTDELIVERY EVENTS HINDER BREASTFEEDING

Interviewees had concerns on the processes that follow after giving birth with some of them noted to be obstacles to early initiation of breastfeeding since time is consumed trying to accomplish them. Routine practices were also seen as hindrances to early initiation of breastfeeding for instance observation of babies with birthweight above four kilograms in the neonatal unit after being born as well as putting newborns in the baby warmer immediately after being born.

### Genital tears repair

participants reported that perineal tear repair process hindered early initiation of breastfeeding since time is consumed in preparing instruments as well as doing the procedure. However, there were suggestions that breastfeeding could still be initiated early despite the perineal tear as long as the mothers and attending nurse are willing. Another aspect of procedure raised was the delay in delivery of placenta requiring manual evacuation, as this was painful and time consuming too.

> *…Even for those who deliver normally, babies might miss the chance to breastfeed immediately, for example, in case a lady had a tear, this can hamper breastfeeding’ (Nurse 1)*
>
> *‘…I believe that nurse would even help me to breastfeed even before starting the tear repair process’ (Mother 3)*
>
> *‘…in my recent delivery, the placenta delayed to get out, and this was painful as if am still about to deliver a baby’ (Mother 1)*

### Routine post-delivery practices

the nurses elaborated the process of care in case a newborn weighs more than 4 kilograms. Close observation of these babies is of paramount importance as they might succumb to hypoglycemia, hence initial feed with formula milk is provided in the neonatal unit. Moreover, some mothers mentioned that babies were routinely placed in the warmer after delivery and this was followed by other activities by nurses hence preventing them from breastfeeding their babies.

> *‘…we provide information that the place for observation will be in the nursery (for big babies) and the baby will be prescribed formula milk (as initial food)’ (Nurse 2)*

### Male presence

the people around after delivery came out as an influencer as some ladies would never breastfeed in front of a man even if it’s the doctor or husband.

> …*Sometimes culture instance, among Chagas’s difficult to breastfeed in front of people you are not close with hence depending on who are the people within labor room, this can be a challenge. (Nurse1)*
>
> *…it’s not possible to breastfeed in presence of a man, even if it’s the husband who is around. (Nurse 3)*

### Dirty surrounding

participants reckoned that the labor room environment after delivery was not encouraging women to start breastfeeding. Some mothers felt like that their bodies were not clean enough after delivery and breastfeeding was postponed until cleanliness has been done.

*‘…you know it’s hard to start breastfeeding in labor room without being comfortable, and after being cleaned, comfortability actually comes, and that is when breastfeeding is possible’ (Mother 2)*

> *…The environment with blood makes mothers hesitate to breastfeed, some of them say that the environment kind reminds them of labor pain, hence for them to relax and breastfeed, they want to be in the ward!! (Nurse 2)*
>
> *…even in delivery room its possible despite some ladies refusing because of the surrounding being shabby sometimes*., *imagine breastfeeding in place full of blood!!! (Nurse 1)*
>
> *‘…hmmm! no! my baby was put on a warmer, and that was it in there’ (Mother 1)*

## NON-CONDUCIVE BREAST-FEEDING ENVIRONMENT

### Lack of care provider support

post-delivery mothers pointed out that lack of support from midwives in initiation of breastfeeding was a barrier to accomplish this process. However, the midwives reported that huge workload contributed to inability to support the mother to initiate breastfeeding early.

> *‘…Sometimes documentation can consume your time, and you may even forget to assist a lady to breastfeed, but this depends on how busy the day has been’ (Nurse 1)*
>
> *…human resource also because some days where there are many deliveries, you might feel you are going nuts hence it’s tough to help mothers start breastfeeding well!! (Nurse 2)*

### Mother’s readiness to breastfeed

Being ready after delivery for breastfeeding came out as an intriguing factor. Mothers reported that being tired and uncomfortable influenced their ability to start breastfeeding and suggested that time is needed for them to be fully ready. There was emergence of a concept that breastfeeding has to occur when post-delivery mother is in good and relaxed mood.

> *…Sometimes mothers are tired after delivery because of labor process hence are not relaxed to start breastfeeding. (Nurse 3)*
>
> *‘…(Alaa!), labor pain hmm!!!!!!you find that a woman is tired after delivery, so it becomes tough to start breastfeeding’ (Mother 5)*
>
> *…I was tired lol, to start breastfeeding within labor room is a big NO’ (Mother1)*

## DISCUSSION

The present study provides insights on the barriers to early initiation of breastfeeding based on experiences among midwives and immediate postpartum mothers as shown in the diagram below:

### Ishikawa diagram highlighting barriers

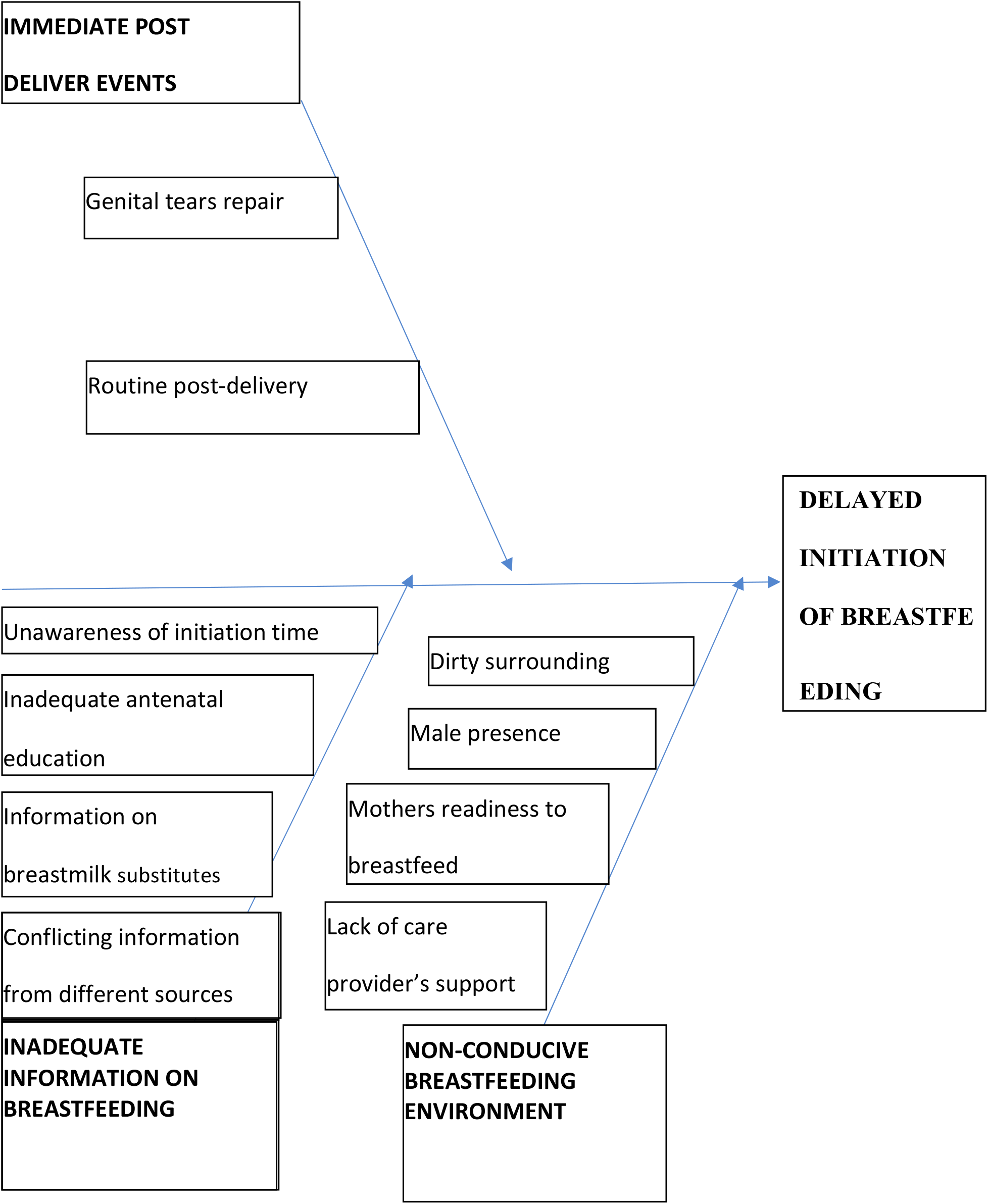

Some of the barriers identified are similar to what has been highlighted in other studies. Information given to pregnant women plays a key role in influencing breastfeeding initiation

In a descriptive cross-sectional study conducted in maternity ward at Mulago hospital in Uganda using mixed methods, inadequate information from ANC was found to impact negatively early initiation of breastfeeding. They suggested that broadening of breastfeeding information to include early initiation is therefore an important addition (39). Another study done in Ethiopia showed that health education and counseling during ANC visits is an intervention that promotes early initiation of breastfeeding since poor attendance of these sessions was seen to be associated with delayed initiation of breast-feeding (18). In a study done in Nepal using data from Demographic and Health Survey from different periods concluded that education and counselling mothers was a successful intervention for proper breastfeeding practices even in challenging situations such as caesarean section deliveries (16).

The Tanzanian antenatal care guideline specifies that breast examination and breastfeeding counseling should be done during the last antenatal care visit. However, specific information about time of initiation of breastfeeding is lacking (40). Therefore, there is a need to broaden the provided information covering early initiation of breastfeeding knowledge among doctors and nurses who attend pregnant mothers at ANC. There is also need to emphasize on the importance of breast examination to identify women who may have challenges in breastfeeding.

Easy availability on information of breast milk substitutes was identified as a barrier in the present study. Similar results have been described from other African countries. A cross sectional study done at maternity ward in Juba Teaching hospital, South Sudan showed that mothers were two times more likely to fail in the practice of early initiation of breastfeeding when they were exposed to breastmilk substitutes advertisement (41) The availability of breastmilk substitutes together with abundant information within our society seems to provide an alternative to breastmilk hence a need to strongly implement measures according to recommendation from WHO code for marketing of breast milk substitutes (15)

An interesting finding from the present study was that some health care providers were not aware of the ideal time to initiate breastfeeding. It’s a basic life principle that you cannot do something without knowing it well. Therefore, initiating breastfeeding without knowing the right timing can be a challenge. In Bangladesh, with support from UNICEF, they have implemented ‘Every Mother Every Newborn Quality Improvement’ initiatives for improving care around the time of childbirth. In this initiative, presence of skilled healthcare provider enhanced early initiation of breastfeeding (42).

Knowledge amongst mothers about the right time for initiation was also poor in our study. Maternal awareness can positively influence breastfeeding since they can work together with care providers during the immediate postpartum period. In a comprehensive cross-sectional health facility-based survey it was revealed that knowing the right time to start breastfeeding among mothers was a positive influencer of early initiation of breastfeeding (43).

Mode of delivery has been seen as an influencer of early initiation of breastfeeding with spontaneous vertex delivery within hospital showing to have positive impact (44). However, reports still shows there are babies who are missing this chance (13). The present study revealed that even after a vaginal delivery, certain practices can negatively impact the time to initiate breastfeeding. These include: procedures such as perineal tear repair, common practices such as placing babies in warmer after delivery or taking babies with weight more than 4 kilograms to the neonatal unit for observation and formula feeding. These practices are not in keeping with recommendations from the BFHIs 10 steps which advocates rooming in and immediate skin to skin contact (23). There is no one agreed plan on caring for newborns who weigh 4 kilograms and above hence principles of rooming in and skin to skin contact should be advocated, however care after delivery should be individualized depending on the babies/mothers’ condition (45). Perineal tear repair timing has been shown to impact initiation of breastfeeding with positive impact when the duration is short (46).

Conduciveness of the environment for initiating breastfeeding emerged as a barrier in the present study. Support from care providers after delivery, body and environmental cleanliness and the surrounding people were mentioned to impact the initiation of breastfeeding. Lack of support from midwives has been shown by other studies to impact negatively early initiation of breastfeeding (46)(47). This lack of support could be due to shortage of staff or a heavy work load which limits the ability of the midwife to provide full support to mother. This was also observed in another Tanzanian study where by shortage of staff were challenges being faced by midwives in an urban setting (48).

Factors such as being unclean, dirty labor room and presence of male personnel were unique barriers which have not been frequently reported in literature. These concerns could be unique to our social and cultural norms and they play a role in limiting early initiation of breastfeeding. Recognition and acceptance of the cultural context in which breastfeeding occurs can help to improve breastfeeding outcomes (49)(50). Concerns about cleanliness can be countered through provision of education that breastfeeding and skin to skin contact is safe soon after delivery even if the mother’s body has not been cleaned.

The fatigue experienced by post-partum mothers limits their ability to start breastfeeding early. A study from Niger reported that nearly half of mothers did not initiate breastfeeding early because of fatigue (51). Presence of a close relative in the labour room can facilitate breastfeeding by supporting the mother who may be tired (52).

## STRENGTHS AND LIMITATIONS

The present study being qualitative in nature describes mothers and midwives’ own experiences on initiation of breastfeeding. Interviews were conducted by one investigator in the local language: Swahili and this omitted the biases that can arise based on language and personalities. The interviews were done within 48 hours of delivery and before hospital discharge and hence the experiences were captured while minimizing recall bias.

This study involved participants whose education level was at least diploma. This may be a limitation since responses from mothers with a lower level of education may be different from what has been identified. In addition, the findings captured in an urban private hospital may be different as compared to a public or rural set up hence posing some challenge in generalization.

## TRUSTWORTHINESS

### Credibility

Multiple data reviews were done. This involved the interpretation of data by the primary researcher and supervisors independently resulting into production of overall categories. Moreover, areas of uncertainty in the transcriptions were cross-checked with respective participants so as to enhance common agreement on understanding of certain points. Cross checking was done through recalling of a specific respondent for clarification of transcribed points for common understanding.

### Transferability

The study setting, participants as well as findings were described to the best of the available information. Being a phenomenological qualitative study, the emerging categories have been stipulated such that a reader can have flexibility in reaching conclusion.

### Dependability

Steps involved in this study were clearly stated, and the encountered limitations had been mentioned. Variability is expected since the present study focused on range of experiences rather than an average on the issues of early initiation of breastfeeding.

### Confirmability

The primary researcher is the senior resident in family medicine with special interest in maternal and child care. The researcher was the one who did the interviews without being judgmental or emotional. He comes from the same Tanzanian culture as all the participants and had no fixed beliefs on the subject of breastfeeding. He did his rotation in the department of obstetrics and gynecology two years ago and was not involved directly in the care of the mothers who were interviewed. During his rotation, he used to work with the midwives under their supervision.

M.A who was methodological supervisor has experience on qualitative studies and has been supervising PhD as well as master’s students in research area. M.N who was a content supervisor is a pediatrician with added expertise as lactational consultant. E.A who was the third supervisor is the senior instructor in family medicine with expertise in qualitative studies in primary care. Interpretation of results was also independently done by the mentioned supervisors till the conclusion of categories and sub categories

## RECOMMENDATIONS

Individualized care keeping in mind individual women’s needs based on their sociocultural practices will foster early initiation. For example, performing breast examination during the antenatal time would help in identifying women who would otherwise have difficulties breastfeeding and facilitate early intervention

Review of guidelines on breastfeeding education during ANC should be done and this must include a clear section on early initiation of breastfeeding. Quality improvement studies should then be done to determine the success of the initiative.

Reviewing hospital policies and practices and ensuring they are evidence based is required to minimize unnecessary delays in breastfeeding. Moreover, prevalence of early initiation of breastfeeding should be incorporated into the key performance indicators for the department.

Further studies involving pregnant women and ANC care providers will give insight into the challenges of providing breastfeeding education during the antenatal period

## CONCLUSION

This study highlights barriers to early initiation of breastfeeding. The key areas identified were related to gaps in knowledge, immediate postpartum practices and individual perceptions of a non-conducive environment.

## Data Availability

Data available upon contacting Library of The Aga Khan University-Tanzania and corresponding author

## ACKNOWLEDGEMENT

The authors are grateful to all the study participants. Special thanks to Dr Munawar Kaguta for his advice and support to the principal investigator.

